# Ethical dilemma arises from optimising interventions for epidemics in heterogeneous populations

**DOI:** 10.1101/2023.02.18.23286135

**Authors:** Pratyush K. Kollepara, Rebecca H. Chisholm, István Z. Kiss, Joel C. Miller

## Abstract

Interventions to mitigate the spread of infectious diseases, while succeeding in their goal, have economic and social costs associated with them. These limit the duration and intensity of the interventions. We study a class of interventions which reduce the reproduction number and find the optimal strength of the intervention which minimises the final epidemic size for an immunity inducing infection. The intervention works by eliminating the overshoot part of an epidemic, and avoids a second-wave of infections. We extend the framework by considering a heterogeneous population and find that the optimal intervention can pose an ethical dilemma for decision and policy makers. This ethical dilemma is shown to be analogous to the trolley problem. We apply this optimisation strategy to real world contact data and case fatality rates from three pandemics to underline the importance of this ethical dilemma in real world scenarios.

## 1 Introduction

Infectious disease epidemics have been suppressed and mitigated using a combination of non-pharmaceutical interventions (NPIs) such as lock downs, social distancing, mask wearing and contact tracing, and by pharmaceutical interventions such as immunising the population using vaccines. In the absence of vaccines, NPIs are the primary option. However, NPIs, and in particular, lock downs, can have significant economic, mental health and social costs associated with them. Instead of protracted or repeated lock downs (as observed during the COVID-19 pandemic), a one-shot intervention has been suggested as a possible alternative for diseases which induce immunity upon recovery from infection. An intense but short-duration lockdown is imposed near the peak of the epidemic to stop the transmission during the overshoot phase of the epidemic and reduce the final size (total number of infections) to the herd immunity threshold of the epidemic (number immune in the population required to stop the growth in infections) [1]. The overshoot phase is when the number of active infections start to decline (effective reproduction number is less than one), but a significant number of new infections are created. The overshoot is the difference between the final size and herd immunity threshold. Thus such interventions reduce the overshoot to zero (See Glossary in the supplementary material for detailed definitions of technical terms).

In this work, we explore an alternative strategy to achieve the same outcome through a prolonged but weaker intervention instead of a short and intense intervention. Such an intervention, if implemented early, will have the added benefit of reducing and delaying the peak of the epidemic as well, in contrast to the one-shot intervention [1]. As with the one-shot intervention, the rationale of this strategy is to calibrate the intervention in such a manner that the final size of the mitigated epidemic is identical to the herd immunity threshold of the original epidemic. Therefore, when the intervention ends, there is no risk of further introductions developing into future epidemics or a second wave of infections. We show that this strategy is an optimal strategy for minimising the final size in the long term.

In the context of COVID-19 modelling, research on optimal interventions has attempted to include economic costs along with the objective of reducing infections: using detailed agent-based models [2] and fine-tuned intervention strategies [3, 4], a balance is sought between socio-economic and health costs to minimise the total cost [5], or the claim that interventions reduce the economic well-being of a society has been challenged [6, 7]. Optimal interventions have also been studied as resource allocation problems where a limited stockpile of vaccine is available or a limited ‘amount’ of social distancing is acceptable and the objective is to find the distribution of the intervention that minimises the reproduction number or a health-related objective function [8, 9, 10].

We do not include economic costs in an explicit manner in our model. The amount of reduction in ℛ_0_ can be interpreted as the cost – the higher the reduction in ℛ_0_, higher the social and economic cost of intervention. The calculations involved in finding the optimal strategy mainly rely on the knowledge of the basic reproduction number (or the next generation matrix). We show that in populations with transmission heterogeneity, implementing an optimal intervention to minimise the final size could involve a moral/ethical dilemma for decision makers, which is analogous to the commonly known trolley problem [11, 12]. The dilemma arises as a result of transmission heterogeneity in the population. We performed a literature search with relevant keywords and were unable to find any research that examined non-pharmaceutical interventions with an ethical dilemma (See supplementary material for keywords). A pre-print, ref. [13], had a similar approach in that they optimised synthetic contact matrices from various European countries to minimise deaths or years of life lost by achieving herd immunity for the COVID-19 epidemics. Their model and intervention scenarios are quite complex: transmission model with six stages of infection, waning immunity and duration of intervention. They find through numerical methods that increasing transmission in younger age groups is required to minimise the years of life lost for the COVID-19 epidemics. Another article, ref. [14], also utilised this strategy of achieving herd immunity in a model with SIR disease and resource growth dynamics in the context of COVID-19. This study did not explore any strategies where transmission is increased. Our work differs from the above-mentioned studies as we explore strategies that increase transmission, in detail and discuss the ethical dilemma. The model we use has the minimal complexity required to explore the underlying mechanisms of this ethical dilemma for a wide range of basic reproduction numbers and to show the impact of heterogeneity and population structure on epidemics and interventions.

In the following sections, we explain the modelling framework, results of our analysis, and conclude with a discussion of our modelling assumptions and the ethical dilemma that decision-makers could face.

## 2 Methods

We used deterministic SIR and SIR-like models to study the optimal intervention. In sub-section 2.1 and 2.2, we explain the models used for a homogeneous population and for a heterogeneous population, respectively, in addition to describing the calculations for finding the optimal intervention. In sub-section 2.3 we describe how this optimisation strategy is applied to real-world data and in sub-section 2.4 we explain how an optimal intervention can be found if there is a delay in the start of the intervention.

### 2.1 Homogeneous population

We use an SIR model with the variables *s, i* and *r* to represent the fractions of individuals in the total population who are susceptible, infected and recovered respectively [15, 16]. The population is assumed to be closed (no entry/exit) and it is normalized such that *s* + *i* + *r* = 1.

In this case, the final size of the epidemic is completely determined by the basic reproduction number *R*_0_ and can be obtained using the following equation [15, 16]:

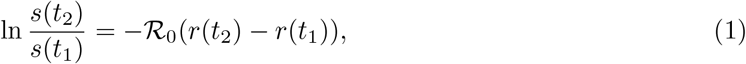

where *s*(*t*_1_) and *s*(*t*_2_) are the fractions of susceptible and *r*(*t*_1_) and *r*(*t*_2_) are the fractions of recovered individuals in the population at time instants, *t*_1_ and *t*_2_. Using the conditions *i*(*t*_1_) *≈* 0, *r*(*t*_1_) = 0 and *i*(*t*_2_) ≈0, which describe the population at the start and end of an epidemic, the well known final size relation can be obtained [17, 15, 16, 18]

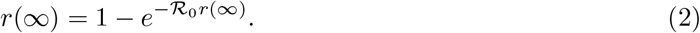

An intervention that reduces transmission would affect the basic reproduction number as ℛ_0_ *→*ℛ _0_(1*−c*) where 0≤*c*≤1. In the case of a homogeneous population, the herd immunity is achieved when the fraction of susceptible individuals in the population is less than 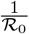. Therefore, we substitute *s*(*t*_2_) = 1*/*ℛ_0_ and *s*(*t*_1_) = 1 and solve for *c*. We find the optimal reduction in the basic reproduction number is

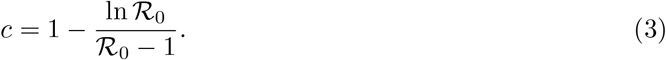

We verify this analytical result in section 3.1 by simulating an epidemic where ℛ_0_ is changed to ℛ_0_(1*−c*) in the early stage of the epidemic and the intervention is switched off once the active infections, *i*(*t*), decline to a negligible number.

### 2.2 Heterogeneous population

In a heterogeneous population, individuals may be further stratified into groups. To represent the fraction of individuals in the total population who belong to a group *k*, we use the variables *s*_*k*_, *i*_*k*_ and *r*_*k*_ such that *s*_*k*_ + *i*_*k*_ + *r*_*k*_ = *n*_*k*_, where *n*_*k*_ is the proportion of the population who belong to group *k* and ∑_*k*_ *n*_*k*_ = 1. Heterogeneity in transmission characteristics can affect the behaviour of epidemics in a significant manner. Epidemics in populations with different transmission structures but identical reproduction numbers can have widely different final sizes. An epidemic in a heterogeneous population can be described by the following SIR-like model, assuming identical duration of infection for all groups and measuring time in the units of the average infection duration,

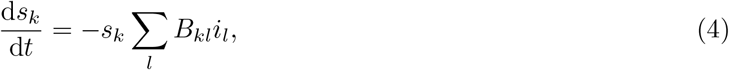

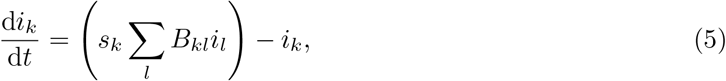

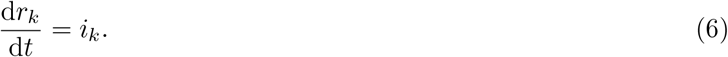

The term *B*_*kl*_ is the average number of infectious contacts that an individual in group *l* causes in group *k*. The next generation matrix, **G** [19], can be constructed for this system with entries:

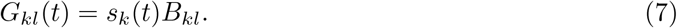

The term *G*_*kl*_ is the expected number of infections that would be caused in fully susceptible group *k* by an infected individual in group *l*. The dominant eigenvalue of **G** gives the reproduction number of the system [19].

The epidemic sizes for this model are given by [17, 18]

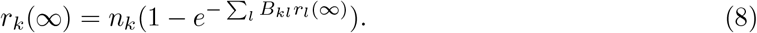

It should be noted that we normalize the final size for the heterogeneous population to be the number of infections in a group as a fraction of the total population.

The recipe for optimisation is similar to the homogeneous case. In the heterogeneous case, we find the level set where the reproduction number is equal to one (analytically in the case of two groups and numerically for more groups), which is the infinite set of values of *s*_*k*_ that would achieve the herd immunity threshold. Then, we optimise subject to this constraint to find the values, 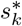, that minimises the cost function (the final size or a weighted sum of final sizes of each group). From this we obtain 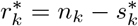. Finding the level set requires finding the proportion of susceptibles of each group, *s*_*k*_, which would ensure that the reproduction number (top eigenvalue of **G**) is equal to one. In equation (8), the final sizes, *r*_*k*_(∞), can be replaced by the optimal final sizes, 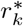, to find the optimal contact matrix. Comparing the original contact matrix with the optimal one tells us how the contact structure of a population must be changed in order to obtain the optimal outcome.

A crucial point to note here is that unlike with the homogeneous population, it is possible for certain elements of **B** and for certain final sizes to increase, 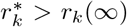, in order to minimise the cost function. In other words, the optimal intervention corresponds to an increase in transmission among certain groups or among pairs of groups. In such cases, the change in reproduction number can not be a measure of the economic or social cost. Nonetheless, this leads to some interesting results which are presented in the next section.

A weighted cost function which is a weighted sum of the final sizes in each group is useful when we are interested in minimising a certain outcome of infections rather than the number of infections, for example, deaths or hospitalisations. The optimisation problem of finding the state of the population which minimises a general objective function and fulfils the herd immunity condition can be solved semi-analytically for the case of two groups and is presented in the supplementary material. For more than two groups, we solve the optimisation problem numerically.

A schematic diagram of the optimisation procedure for both homogeneous and heterogeneous population is shown in Supplementary Figure 6.

### 2.3 Real world contact matrix

We used a contact matrix calculated using surveys from a sample population stratified into six age groups in the Netherlands [20]. The contact matrix scaled by a disease specific parameter gives the next generation matrix. Using the next generation matrix and the age distribution, the optimal intervention for a given cost function can be obtained. We calculated the optimal intervention using this contact matrix for a range of ℛ_0_ values and four different cost function weightings (an unbiased cost function and three from observed case fatality rates (CFRs) of 2009 pandemic in Mexico and 1918 pandemic in the USA and COVID-19 pandemic) [21, 22, 23]. The age groups, their population sizes and CFRs are shown in Table 1. Note that the age stratification used in the CFR study for the 1918 pandemic do not match exactly with the age groups of the contact matrix and furthermore the estimates were extracted from figures. For the 2009 pandemic and COVID-19, CFR was reported with a high age resolution but the size of the age groups was not immediately available. Due to lack of data on infection fatality rates, we are using the CFRs as a proxy for the probability that an infected individual dies. Thus the CFR values and the results relying on them are meant to be for illustration purposes only.

**Table 1:** The age groups used in the contact matrix from ref. [20], their names used in this article, size of the group (as a proportion of the total population), approximate estimates of case fatality rates (CFRs) obtained from refs. [21, 22, 23].

| Age group (years) | Group name | Group size (Netherlands) | CFR (COVID-19/ multinational) | CFR (2009 flu/ Mexico) | CFR (1918 flu/ USA) |
| --- | --- | --- | --- | --- | --- |
| 1-5 | Children | 0.06 | 0.000031 | 0.0096 | 0.0168 |
| 6-12 | Pre-adolescents | 0.09 | 0.000008 | 0.0038 | 0.0047 |
| 13-19 | Adolescents | 0.11 | 0.000028 | 0.004 | 0.0082 |
| 20-39 | Young adults | 0.34 | 0.000214 | 0.026 | 0.0272 |
| 40-59 | Middle adults | 0.23 | 0.001807 | 0.054 | 0.0149 |
| 60 + | Elderly | 0.17 | 0.029520 | 0.059 | 0.0378 |

The severity of the dilemma in the optimal intervention can be quantified through the number of infections (or deaths) caused due to the intervention per infection (or death) prevented. It can be calculated using

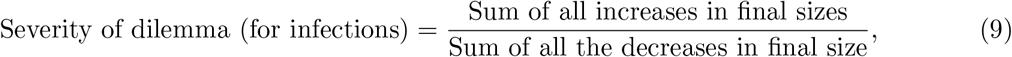

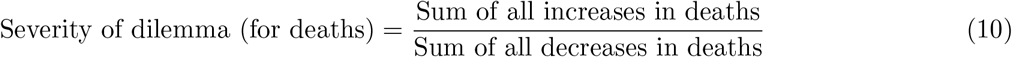

### 2.4 Delayed intervention in a homogeneous population

In the above sections, we have assumed that the basic reproduction (or the next generation matrix) is a known entity and therefore an intervention is implemented right at the start of the epidemic. Calculating the strength of the optimal intervention requires knowledge of the reproduction number, the intervention would have to start after the epidemic has been established and enough observational data has been collected to calculate the reproduction number. While the basic principle would still hold, a delay could change the strength of the optimal intervention. To find the optimal strength for a delayed intervention, we use the final size relation with the final state *s* = 1*/*ℛ_0_, *i* = 0 and an initial arbitrary state *s*_*L*_, *i*_*L*_ at a time instant *t*_*L*_ when the intervention begins. We replace basic reproduction number in equation (1) with ℛ_0_(1 *− c*) and solve for

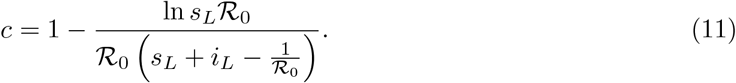

Using a numerical solution of the SIR equations, *s*_*L*_ and *i*_*L*_ can be found and the above equation can be solved for *c*. The equation (11) reduces to equation (3) when *s*_*L*_ = 1, *i*_*L*_ = 0, and *c* = 1 when *s*_*L*_ = 1*/*ℛ_0_ and *i*_*L*_ *>* 0. If 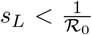, then *c >* 1 which is biologically meaningless and reflects the fact that the population is already below the herd immunity threshold.

### 2.5 Model assumptions

The homogeneous model assumes that all individuals in the population are identical and every individual is equally likely to come in contact with every other individual. To introduce some complexity in this model, we use the heterogeneous model where individuals are stratified into homogeneous groups. We are using deterministic differential equation models with continuous variables to simulate the dynamics. This means that the number of active infections can decay exponentially but can never reach zero. Thus, the models used here can not simulate a scenario in which an intervention eliminates a disease before reaching herd immunity threshold, as was the case in Australia, New Zealand, Hong Kong, mainland China, Singapore and several other jurisdictions (broadly known as the Zero-COVID strategy). Throughout the paper, we use the Susceptible→ Infected→ Recovered (SIR) disease progression. Therefore, our analysis applies to diseases that induce long term immunity or for which re-infection is not possible.

## 3 Results

### 3.1 Homogeneous population

Simulation of the SIR model differential equations confirms our assertion in equation (3). As shown in Figure 1, a ‘weak’ intervention reduces the final size but does not reduce the overshoot to zero. A strong intervention, on the other hand, reduces the final size as long as the intervention is in place but a resurgence occurs as soon as the intervention ends. The final health outcome under the strong intervention is worse than (or at least comparable to) the weak intervention, while incurring a higher social and economic cost during the intervention. The resurgence occurs because the small number of infections and sufficient number of susceptibles remaining in the population lead to new infections after the intervention is lifted. An intervention which is strong enough to minimise the final number of infections, while avoiding a resurgence, is the one whose final size (during the intervention) matches the herd immunity threshold of the unmitigated epidemic. This is the optimal intervention.

**Figure 1:**
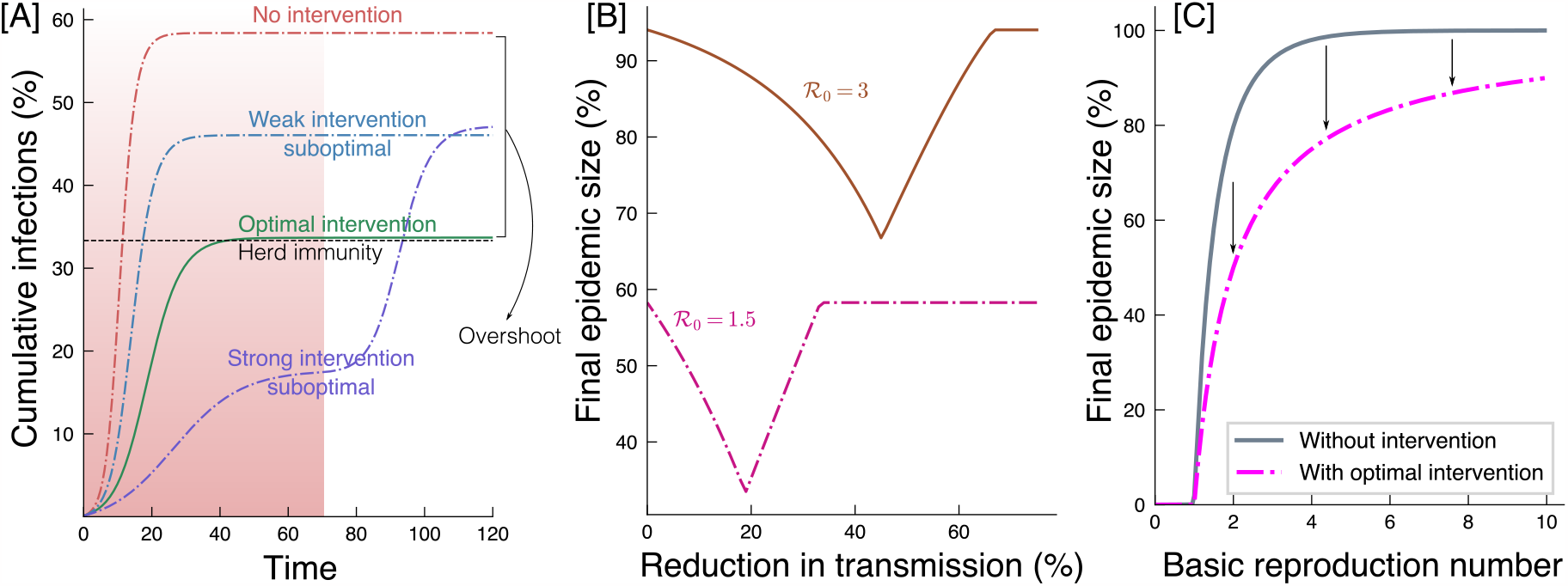
(A) Simulation of four types of intervention for an epidemic with ℛ_0_ = 1.5 in a homogeneous population: (i) No intervention – leads to largest final size, (ii) Weak intervention – reduces the final size, (iii) Strong intervention – reduces final size during the intervention, but leads to a resurgence in infections once the intervention is removed, (iv) A moderate intervention but optimal – final size is same as the herd immunity threshold. (B) The global minima for the final size shows that an optimal intervention strength exists. The resurgence of infections under a strong sub-optimal intervention is subject to certain assumptions which are discussed in the text. (C) The final size without any intervention (equation 2) and the final size with optimal intervention (same as the herd immunity threshold for Recovered state) is shown against the basic reproduction number.

### 3.2 Heterogeneous population

Introducing heterogeneity in the model opens up a space of interventions that is not seen in the homogeneous case. In the homogeneous case, the herd immunity threshold is defined by a single point, but in the case of a structured population, the threshold is given by a collection of points. This can be seen by considering the following: the condition required for reaching herd immunity is that the typical infected individual must not infect more than one individual. In the homogeneous case, one can randomly choose a sufficient number of individuals and immunise them to ensure that the number of infectious contacts is less than one. If the population is structured, the typical infected individual must not infect more than one individual, on an average. As long as the average number of infectious contacts is less than one, herd immunity is achieved, irrespective of how the immunisation has been distributed among the various groups in the population. Thus there are infinitely many interventions that lead to herd immunity and prevent resurgence. Out of all these possibilities, we define the optimal intervention to be the one which minimises the final size.

When the population can be described using two sub-populations or groups, the optimal intervention belongs to one of the following types: the first group is fully infected, the second group is fully infected, or none of the groups are fully infected. This creates the possibility that under the optimal intervention, the number of infections in one of the groups is larger than what would have occurred in the unmitigated epidemic, subject to the structure of the population. In Figure 2 we show an example in which this occurs. This leads to an ethical dilemma wherein a certain group in the population incurs a higher cost (due to an increased number of infections) than would have happened without the intervention in order to minimise the cost for the whole population. Thus, the non linearity of the infectious disease dynamics, combined with population structure, lead to an ethical dilemma for policy/decision-makers which is analogous to the well-known trolley problem [11, 12] (see Figure 2). The trolley problem involves a setup in which a train is going to hit a group of people who are lying on the tracks. The train can not be stopped, but a lever can be pulled to switch the train onto a different track on which fewer people are lying. The dilemma that is posed by this situation is whether it is ethical to save more lives by ending a lesser number of, but, different lives?

**Figure 2:**
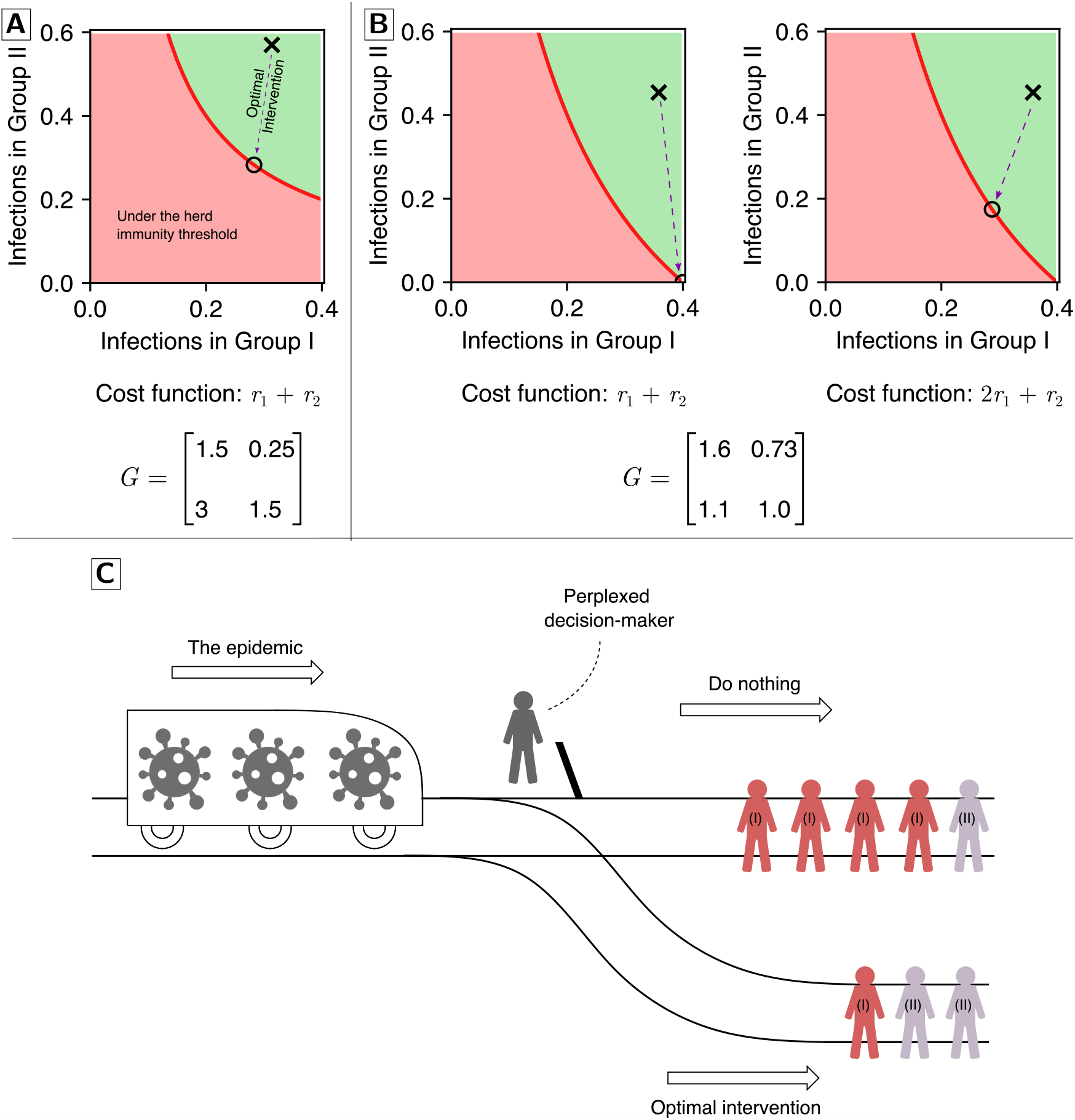
Population structure and non linearity of infectious disease dynamics lead to an ethical dilemma. (**A**) A plot of the infections in the two groups of the population. The corresponding cost function and next generation matrix are shown. The red curve shows the herd immunity threshold, the cross shows the final size without an intervention, the circle shows the final size if the optimal intervention is used. (**B**) A comparison of the interventions when the cost functions are different. The first plot shows that the optimal intervention leads to an increase in the number of infections in the first group. It is an example of the ethical dilemma of implementing the optimal intervention, which is explained further in (**C**). The second figure shows the plot for the cost function when the first group is given twice the weight as the second group, which means prevention of infections in the first group takes precedence over the second group. In this case, the intervention reduces the infections in both the groups. (**C**) The ethical dilemma involved in implementing optimal interventions is analogous to the well-known ‘trolley problem’. If the decision-maker does not act, the incoming epidemic (represented as a trolley) is going to cause many infections. If the decision-maker implements an optimal intervention (switches the tracks), the number of infections in the total population is minimised, but someone who otherwise would have been safe from infection, becomes infected (shown by the increased number of infections of group (II)).

Diseases often lead to a worse health outcome (mortality rate, hospitalisation rate, chance of leading to chronic conditions etc.) in certain groups of the population (the elderly age groups for instance). Instead of minimising the final size of the epidemic (which is the sum of final sizes in each group), it may be more prudent to minimise a cost function which is a linear combination of the final sizes in the groups, such that a group with a worse outcome of infection is given a higher weight in the cost function. As changing the cost function would change the optimal solution, the cost function plays a role in determining the ethical dilemma. For the example shown in Figure 2, the ethical dilemma is no longer present if a weighted cost function is used as infections are being reduced in both groups.

### 3.3 Real world contact matrix

When the basic reproduction number is close to one, at least one of the age groups is required to endure a higher final size for all the cost functions we used (Figure 3 and Supplementary Figures 3, 4 and 5). The cost functions are weighted using estimates of case fatality rates (CFRs) of the 2009 influenza pandemic, the 1918 influenza pandemic, and COVID-19 pandemic [21, 22, 23]. In addition to looking at the final sizes and how they change in various age groups, we can use the case fatality rates to estimate the deaths in each of the age groups and how they change with the optimal intervention.

**Figure 3:**
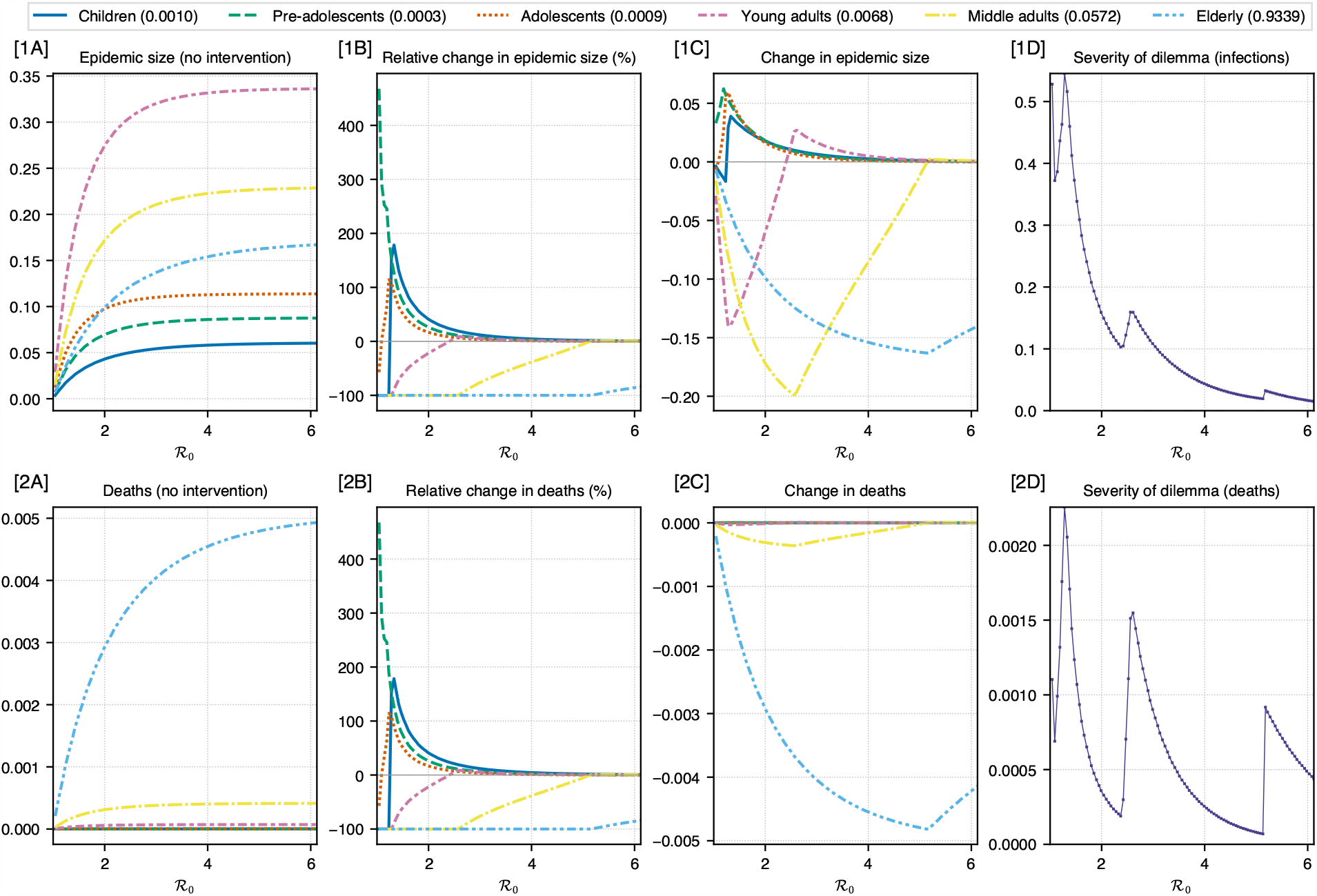
COVID-19 pandemic: A real-world contact matrix from a sample of the Dutch population is used to determine the effect of optimal intervention on different age groups for a range of ℛ_0_ values. Estimates of case fatality rates for the COVID-19 pandemic in Mexico have been used to weight the cost function (for minimising total deaths in the population) [23]. Rows (1) and (2) show the plots for infections and deaths respectively. Column (A) shows the epidemic size and deaths if no intervention was performed. Column (B) shows the relative change in epidemic size and deaths under optimal intervention. Column (C) shows the magnitude of change in epidemic size and deaths under optimal intervention. Column (D) shows the severity of the ethical dilemma (see main text for definition) with ℛ_0_. The legend for columns (A, B, C) shows age groups and the number in bracket shows the weight assigned to it in the cost function. These weights are proportional to the case fatality rates.

For the COVID-19 pandemic, pre-adolescents have the lowest CFR, and it increases for higher age groups (Figure 3 and Table 1). Figure 3 (columns B and C) shows that as ℛ_0_ is increased, the age groups start to experience an increase in infections (relative to no intervention case) in the following order – pre-adolescents, adolescents, children and finally young adults, which is also the order in which the CFR increases. Thus, the CFR may explain the nature of the ethical dilemma. At ℛ_0_ *≈*1.3, the severity of ethical dilemma is highest, with 0.55 new infections for every infection prevented and 0.0025 new deaths for every death prevented (panels 1D and 2D of Figure 3). The severity for deaths is quite low compared to other pandemics because of the large disparity in the CFR across age for COVID-19.

For the 2009 flu pandemic (Supplementary Figure 4), pre-adolescents have the lowest CFR, and it increases with age (Supplementary Figure 4 and Table 1). Supplementary Figure 4 (columns B and C) shows that as ℛ_0_ is increased, the age groups start to experience an increase in infections (relative to no intervention case) in the following order – adolescents, pre-adolescents, and finally young adults, which is not in the increasing order of CFRs. Thus, the CFR does not explain the nature of the ethical dilemma. For infections, the severity of dilemma is highest at about ℛ_0_ = 1.3, where 0.44 new infections are created for every infection prevented. For deaths, the dilemma is the most severe at both ℛ_0_ = 1.3 and 2.25, where 0.045 new deaths are caused for every death prevented.

For the 1918 flu, the CFR with age is often described as a ‘W’ shaped curve (Supplementary Figure 5 and Table 1). Supplementary Figure 5 (columns B and C) shows that as ℛ_0_ is increased, the age groups start to experience an increase in infections (relative to no intervention case) in the following order – adolescents, pre-adolescents, and finally middle adults, which is also the order in which the CFR increases. Thus, the CFR may explain the nature of the ethical dilemma. At the peak of severity (ℛ_0_*≈*1.3), 0.47 new infections are created for each infection prevented and 0.12 new deaths for every death prevented.

For realistic CFRs, the dilemma in terms of infections is quite severe at its worst, with almost 1 person getting infected for protecting two individuals from getting infected. Some features of the ethical dilemma are common to all three pandemics. As ℛ_0_ is increased, the severity of dilemma never quite reaches zero but seems to approach zero in a non-monotonic manner and pre-adolescents and adolescents always endure an increase in infections. The nature of the ethical dilemma may be explained by the CFRs of the age-groups in the 1918 and COVID-19 case, but not in the case of the 2009 pandemic.

For an unbiased cost function (Supplementary Figure 3), we see very different results. The severity of dilemma is zero for most of the ℛ_0_ range. In the space where the dilemma does occur, only the young adults and adolescents experience an increase in final size. At the most severe ethical dilemma, 0.175 new infections are created for every infection prevented.

### 3.4 Delayed intervention

We calculate the optimal strength of the intervention and simulate the model to confirm the mathematical analysis in Section 2.4. Using equation (11) we observe that the strength of optimal intervention increases in a super-linear manner with the duration of delay. The results are presented in Figure 4. As the population approaches the herd-immunity threshold, the strength of intervention approaches one – corresponding to the one-shot intervention [1].

**Figure 4:**
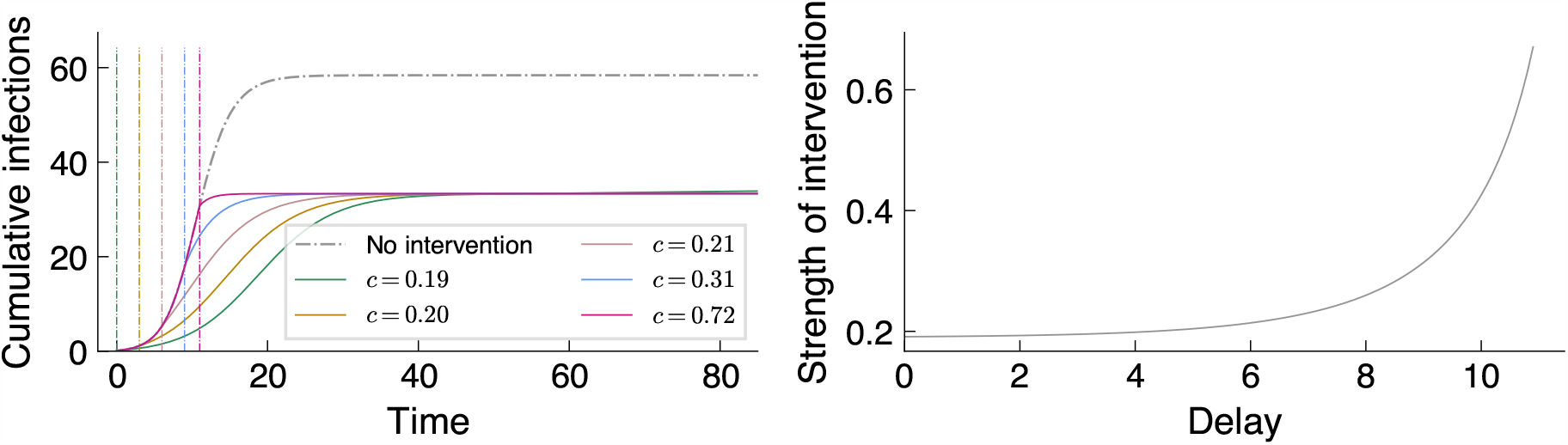
**Left:** The time series of cumulative infections (*i* + *r*) for optimally controlled epidemics for a range of delays in the intervention. The vertical dash-dot lines show the time at which the intervention starts, and its corresponding time series is represented by the same color. **Right:** The strength of optimal intervention, *c*, plotted against the delay in intervention *t*_*L*_ using equation (11). The super-linear increase in the strength, *c*, shows the need for an early implementation. **Parameters:** Homogeneous SIR model with basic reproduction number ℛ_0_ = 1.5 and *γ* = 1. Intervention is implemented for a duration of 50 time units.

## 4 Discussion

In this work, we have examined a strategy of optimal intervention which allows the epidemic to cause just enough infections to induce herd immunity, eliminate the overshoot, and prevent future introductions from becoming epidemics. In addition to minimising the final size, this intervention would also slow down the growth of the epidemic and reduce the peak, which allows time to develop treatments and increase healthcare capacity. For a homogeneous population, the results are straight-forward – decrease the transmission by a pre-determined amount so that the final size reaches the herd immunity threshold and no more. A sensitivity analysis of the homogeneous model and intervention strategy was performed where the optimal strength of intervention and the resulting final size were computed for both the actual value of ℛ_0_ and a ‘measured’ value of ℛ_0_ with four different error rates (See Supplementary Figure 2). We find that in both underestimation and overestimation of ℛ_0_, the epidemic size is larger, but it is better to overestimate ℛ_0_.

In the case of heterogeneous transmission, our results indicate that the optimal strategy may require increasing infection in some of the groups and decreasing it in others, in order to minimise the final size for the whole population. This is analogous to the trolley problem, and it calls for a discussion around the ethics of subjecting certain groups to a higher rate of disease incidence, and the feasibility of this policy. If increasing transmission in certain groups is not viable either due to operational reasons or ethical considerations, herd immunity can still be achieved (and resurgence prevented) by reducing transmission in all groups. We have also explored the role of the cost function in determining the ethical dilemma by weighting the final sizes of different age groups using case fatality rates of the 1918, 2009 and COVID-19 pandemic and shown that the ethical dilemma happens in all three cases. Our work shows that even without an explicit consideration of economic and social costs of an intervention, there are challenging ethical questions to be answered for the first order problem of minimising the final size.

The optimal interventions shown for the 1918, 2009, and COVID-19 pandemics are not meant to be policy advice because the estimates of CFR were approximate and also because influenza and COVID-19 can be described by an SIR-like model only when new variants of the pathogen do not emerge. They are meant to show that the ethical dilemma we have discussed in this paper is not merely a theoretical observation in the parameter space of the mathematical model but a possibility that one should be aware of for future epidemics and pandemics.

Ethical dilemmas in public health are well known, and there have been debates on prioritization based on age for vaccination during the COVID-19 pandemic [24]. Prioritising one age group means that another group receives less protection, but under no circumstances does a discriminatory vaccine distribution policy increase the chances of infection in a group compared to the no vaccine scenario. Thus the dilemma presented in our paper is fundamentally different to a vaccine allocation dilemma and is equivalent to the trolley problem. We have used mathematical modelling to show that optimal interventions may require a policy-maker to contend with a trolley problem like situation where the epidemic under an optimal intervention will infect someone who would not have been affected if there was no intervention. We found two works which use an intervention similar to ours, but they did not consider increase in transmission as a strategy for optimal outcomes [14] or did not discuss the ethical implications [13]. Therefore, we believe that our paper contributes to an important point of discussion with regards to optimality of non-pharmaceutical interventions.

In addition to the ethical dilemma shown through our modelling here, interventions which require increasing transmission prompt an ethical discussion in relation to disadvantaged groups. Cultural, economic and social conditions factor into the contact structure of any human population – a high number of contacts due to living in close spaces, a high susceptibility to infection due to preexisting health conditions, or poor access to healthcare facilities, etc. Mathematical models of epidemics can throw light on possible choices of policy and may even help us pick the ones that lead to optimal outcomes. But the decisions made by policymakers are intertwined with political will, their popularity, and social attitudes. These eventually determine whether a particular intervention is favoured by a decision-making body [25, 26]. Disadvantaged groups, across the world, do not exercise sufficient political power to represent their interests in decision making bodies. In such a case, a decision-making body may find it convenient to subject a disadvantaged group to a higher final size in order to decrease the net final size for the whole population and achieve herd immunity. The intervention strategy presented here, always carries such risks with it; and representation of disadvantaged groups thus becomes essential, especially for a policy such as this one.

There are also some practical limitations to the strategy presented here. There would be a natural tendency for individuals to protect themselves from getting infected even if interventions are not in place, so asking individuals to increase their transmission may not be a feasible strategy [27]. The optimal interventions could require a group of individuals to fully isolate themselves from the rest of the population. Such interventions are difficult to implement, as there would always be a small possibility for infections to be introduced into the isolated group [27]. If the transmission in other groups is increased, it would imply a larger chance of introduction into the isolated group.

We have assumed an SIR structure for disease progression in an individual. But, as long as the disease can be reasonably described by a model in which individuals do not become susceptible after getting infected, we would expect our results to be valid. A crucial detail that we have ignored is the stochastic and discrete nature of disease spread since it can capture the elimination behaviour of outbreaks, i.e., it can incorporate the difference between existence and absence of infections. The deterministic assumption and the use of continuous variables in our model means that after an intervention is over, the small number of infections present in the population will lead to another epidemic if herd immunity is not achieved (shown in Figure 1, strong intervention). This however, is one of the possible outcomes. It is possible that the intervention completely eliminates all infections in the population, in which case a new epidemic does not result from any residual infections. However, even in this case the population remains vulnerable to an epidemic due to lack of herd immunity. Thus, a new epidemic can occur if new infectious individuals are introduced into the population. Another possibility is that the epidemic may get established with a delay due to the stochastic dynamics. Factors around contact-tracing and surveillance capacity (to eliminate the disease) and travel restrictions (to prevent introduction of new infections) are important for the selecting the optimal policy response, in addition to the results presented here.

## Data Availability

All data produced in the present work are contained in the manuscript. Software used for analysis is available at https://github.com/Joel-Miller-Lab/optimal-intervention DOI: 10.5281/zenodo.10386309.

## Ethics

Ethics approval was not required for this study.

## Author Contributions

PKK: Conceptualization, Formal analysis, Investigation, Methodology, Software, Visualization, Writing — original draft, Writing — review and editing

RHC: Conceptualization, Investigation, Methodology, Project administration, Supervision, Writing — review and editing

IZK: Conceptualization, Methodology, Supervision, Writing — review and editing

JCM: Conceptualization, Investigation, Methodology, Project administration, Supervision, Writing — review and editing

## Acknowledgements

We thank Michael Meehan, Sarah Becirevic and Somya Mehra for helpful discussions.

## Data Access

The software used in this paper is available at https://github.com/Joel-Miller-Lab/optimal-intervention.

