## Supplemental Information for "Ethical dilemma arises from optimising interventions for epidemics in heterogeneous populations"

Pratyush K. Kollepara[1], Rebecca H. Chisholm[1,2], István Z. Kiss[3], and Joel C. Miller[1]
[1] *Department of Mathematical and Physical Sciences, La Trobe University, Melbourne, Australia*
[2] *Melbourne School of Population and Global Health, The University of Melbourne, Melbourne, Australia and*
[3] *Network Science Institute, Northeastern University London, London E1W 1LP, United Kingdom*

### SUPPLEMENTARY MATERIAL

#### A. Glossary of terms

1. Basic reproduction number ($\mathcal{R}_0$): It is the expected number of new infections created by a typical infected individual when the population is almost susceptible.

2. Effective reproduction number: Same as the basic reproduction number, without the requirement that the population be almost susceptible.

3. Final size of the epidemic, final size, or epidemic size: The total number of infections caused during the epidemic expressed as a proportion of the total population.

4. Herd immunity threshold: The proportion of the population that needs to be immune in order for the growth rate of the epidemic to be zero. This is usually expressed as the proportion of the total population that is immune or recovered (in an SIR model).

5. Overshoot: The difference between the final size and the herd immunity threshold is called the overshoot.

6. Next generation matrix: A generalisation of the reproduction number, useful for populations with heterogeneity or structured populations. Each element of this matrix is a reproduction number between a pair of groups i.e. the element $G_{kl}$ of a matrix $\mathbf{G}$ is the number of new infections in group $k$ created by an individual in group $l$. The reproduction number is the highest eigenvalue of the next generation matrix.

#### B. Literature Search

For the literature search, we used the following query: `(epidemic) AND (intervention) AND (dilemma OR ethical OR ethics OR moral)`, on Web of Science and searched all collections (`https://www.webofscience.com/wos/alldb/summary/9031be14-3217-4085-b901-e04696f4ceb6-9ffdcfc6/relevance/1`). It returned 973 results sorted by relevance and we browsed through the abstracts of first 100 results and did not find any entries that seemed similar to the results presented in our work.

#### C. Finding the optimal intervention

The generalised cost function depends on the weight vector $\mathbf{a} = (a_1, a_2, \dots)$ and the attack rate vector $\mathbf{r} = (r_1, r_2, \dots)$ and is given by:

$$C(\mathbf{r}, \mathbf{a}) = \mathbf{a} \cdot \mathbf{r} = \sum_k a_k r_k. \tag{1}$$

The vector $\mathbf{r}$ is constrained such that $\rho(\mathbf{G}) = 1$, where $\mathbf{G}$ is the next generation matrix and $\rho(\cdot)$ is the dominant eigenvalue.

We present a semi-analytical solution to this optimisation problem when there are two groups in the population. Minimising the cost function $C$ is equivalent to maximising the function $f$ :

$$f(s_1, s_2, a_1, a_2) = a_1 s_1 + a_2 s_2, \tag{2}$$

while the constraint is given by equating the eigenvalue of the next generation matrix to one

$$g(\mathbf{G}) = 1 - \operatorname{Tr}\mathbf{G} + \det \mathbf{G} = 0, \tag{3}$$

$$= 1 - (s_1 B_{11} + s_2 B_{22}) + s_1 s_2 \Delta = 0, \tag{4}$$

where $\Delta = \det \mathbf{B}$. Values of $s_1$ and $s_2$ that satisfy this equation may correspond to cases where the non-dominant eigenvalue is unity. Therefore, the dominant eigenvalue needs to be computed numerically in order to impose this restriction.

To solve this optimisation problem we define a new function $h = f + \lambda g$, where $\lambda$ is a Lagrange multiplier. The partial derivatives of $h$ with respect to $s_1$, $s_2$ and $\lambda$ will be zero at points of interest and the three equations will be solved simultaneously to obtain

$$\frac{1}{\lambda} = \pm\sqrt{\frac{B_{12}B_{21}}{a_1 a_2}}, \tag{5}$$

$$s_1^* = \frac{B_{22} \pm \sqrt{\frac{B_{12}B_{21}a_2}{a_1}}}{\Delta}, \tag{6}$$

$$s_2^* = \frac{B_{11} \pm \sqrt{\frac{B_{12}B_{21}a_1}{a_2}}}{\Delta}. \tag{7}$$

For a given matrix $\mathbf{B}$, group sizes $n_1$ and $n_2$, and costs $a_1$ and $a_2$, the function $f$ will be evaluated for the two points given by $(s_1^*, s_2^*)$ and the four boundary points. The six ‘types’ of possible extrema for the optimisation problem, are shown in TABLE I and in FIG. 1. Depending on the parameters, one of these points will maximise the function $f$ subject to the constraint $g = 0$.

The boundary extremal points represent scenarios where one of the groups is either fully infected or is fully protected from getting infected. Due to the large number (six) of parameters governing the optimal solution, we present graphical representations of the outcomes under the optimal intervention, in each group for sections of the parameter space in FIG. 1. We use a transmission matrix with a symmetric structure $B_{kl} = b_k b_l((1-\alpha)\delta_{kl} + \alpha)$ where $\alpha$ can be used to control the relative magnitudes of the diagonal and off-diagonal elements. The changes in the type of solution occur primarily based on the validity of the extremal points ($r_1^* \in [0, n_1]$ and $r_2^* \in [0, n_2]$) and which of the valid solutions minimise the cost. As an example, the straight line separation between type 5 (local minima) and type 2 (boundary point) and between type 5 and type 0 (boundary point), in the first

TABLE I. List of possible extremal points for a two-group population. The first column shows the solution type referred to in FIG. 1. The second and third column give the expression for the extremal values of $s_1$ and $s_2$. The quantities $B_{kl}$ are the elements of the transmission matrix, $\{a_1, a_2\}$ are the cost function weights. The quantity $\Delta = B_{11}B_{22} - B_{12}B_{21}$.

| Type | $s_1^*$ | $s_2^*$ | Description |
|---|---|---|---|
| 0 | $0$ | $\frac{1}{B_{22}}$ | Boundary point: Group 1 fully infected |
| 1 | $n_1$ | $\frac{1-n_1B_{11}}{B_{22}-n_1\Delta}$ | Boundary point: Group 1 fully protected |
| 2 | $\frac{1}{B_{11}}$ | $0$ | Boundary point: Group 2 fully infected |
| 3 | $\frac{1-n_2B_{22}}{B_{11}-n_2\Delta}$ | $n_2$ | Boundary point: Group 2 fully protected |
| 4 | $\frac{B_{22}+\sqrt{B_{12}B_{21}a_2/a_1}}{\Delta}$ | $\frac{B_{11}+\sqrt{B_{12}B_{21}a_1/a_2}}{\Delta}$ | Local extrema |
| 5 | $\frac{B_{22}-\sqrt{B_{12}B_{21}a_2/a_1}}{\Delta}$ | $\frac{B_{11}-\sqrt{B_{12}B_{21}a_1/a_2}}{\Delta}$ | Local extrema |

row of FIG. 1, can be explained by imposing $s_1^* \geq 0$ and $s_2^* \geq 0$ on the expressions listed in TABLE I.

### D. Additional Figures

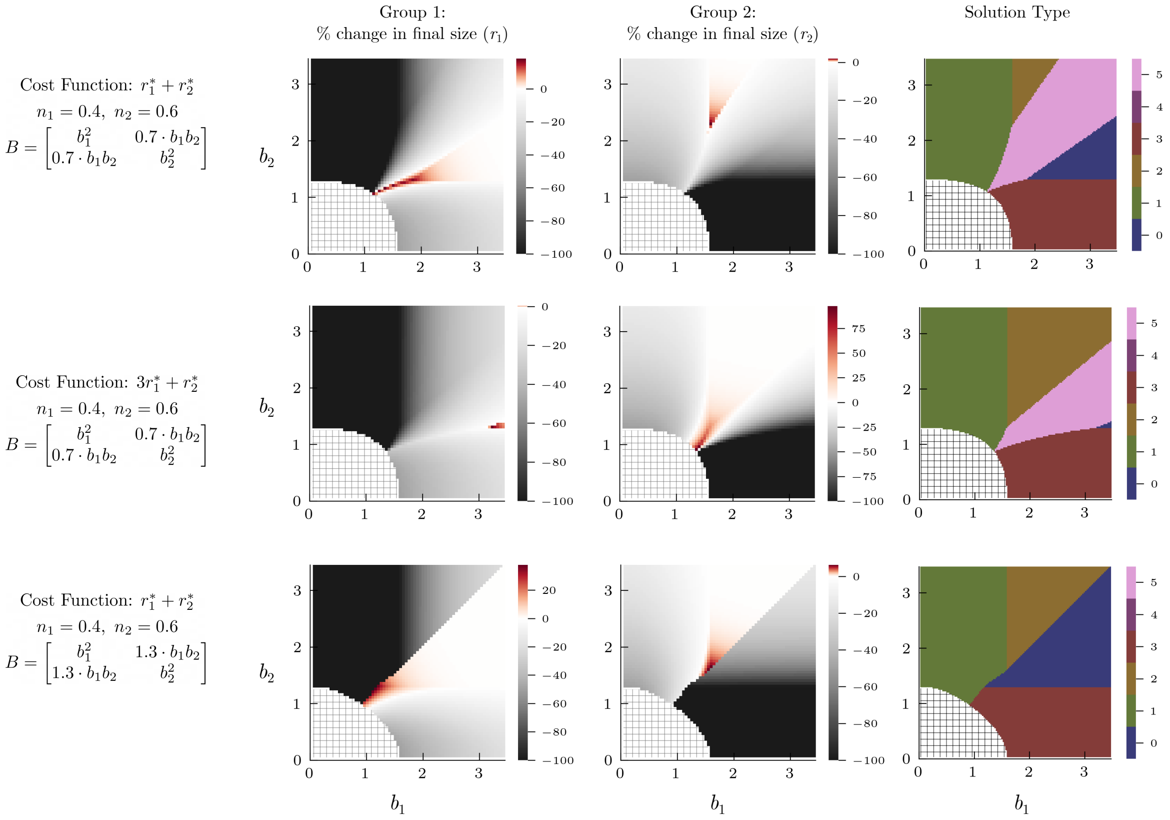


FIG. 1. A section of the parameter and optimal outcome space: The first column of each row shows the cost function, group sizes and the structure of the transmission matrix $B$. The next two columns are heat-maps which show the percentage change in the attack rate of Group 1 and 2 if the optimal intervention is implemented. The last column shows the type of solution obtained from the optimisation problem – the mathematical expression and description for the solutions can be looked up in TABLE I. The red coloring in the heat-map show regions of the parameter space where the optimal intervention would cause a worse outcome in one of the groups while minimising the cost function for the overall population. The model is identical for the first two rows, but the cost function is different. In the first row the cost function is the attack rate, while in the second row it is a weighted function of the two attack rates. The group sizes are 0.4 and 0.6 for all the rows.

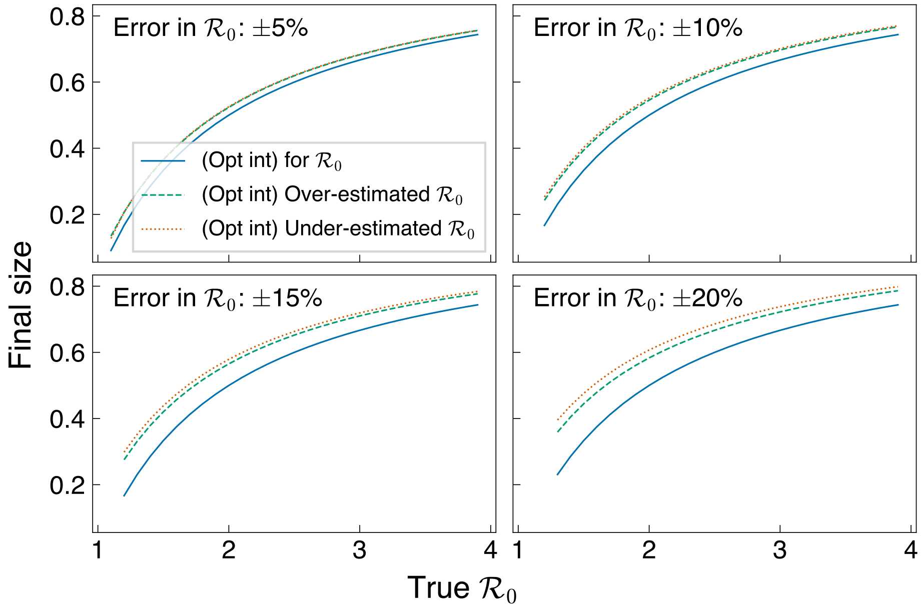


FIG. 2. Error analysis for optimal intervention in homogeneous population: X-axis shows the true value of basic reproduction number and each subplot shows the final size for the true value and with a positive and negative error rate.

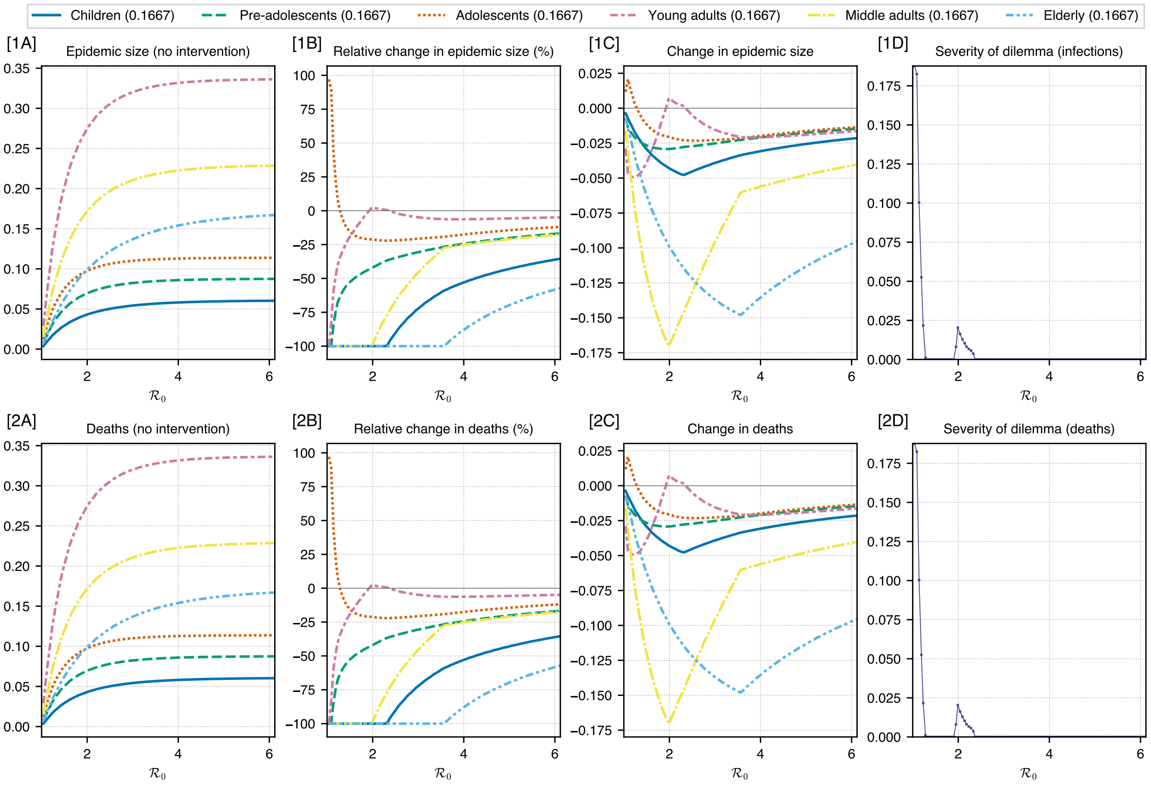


FIG. 3. Unbiased cost function: A real-world contact matrix from a sample of the Dutch population is used to determine the effect of optimal intervention on different age groups for a range of $\mathcal{R}_0$ values. The cost function is unbiased, as in, it is the sum of infections in all groups. Rows (1) and (2) show the plots for infections and deaths respectively. Column (A) shows the epidemic size and deaths if no intervention was performed. Column (B) shows the relative change in epidemic size and deaths under optimal intervention. Column (C) shows the magnitude of change in epidemic size and deaths under optimal intervention. Column (D) shows the severity of the ethical dilemma (see main text for definition) with $\mathcal{R}_0$. The legend for columns (A, B, C) shows age groups and the number in bracket shows the weight assigned to it in the cost function. These weights are proportional to the case fatality rates.

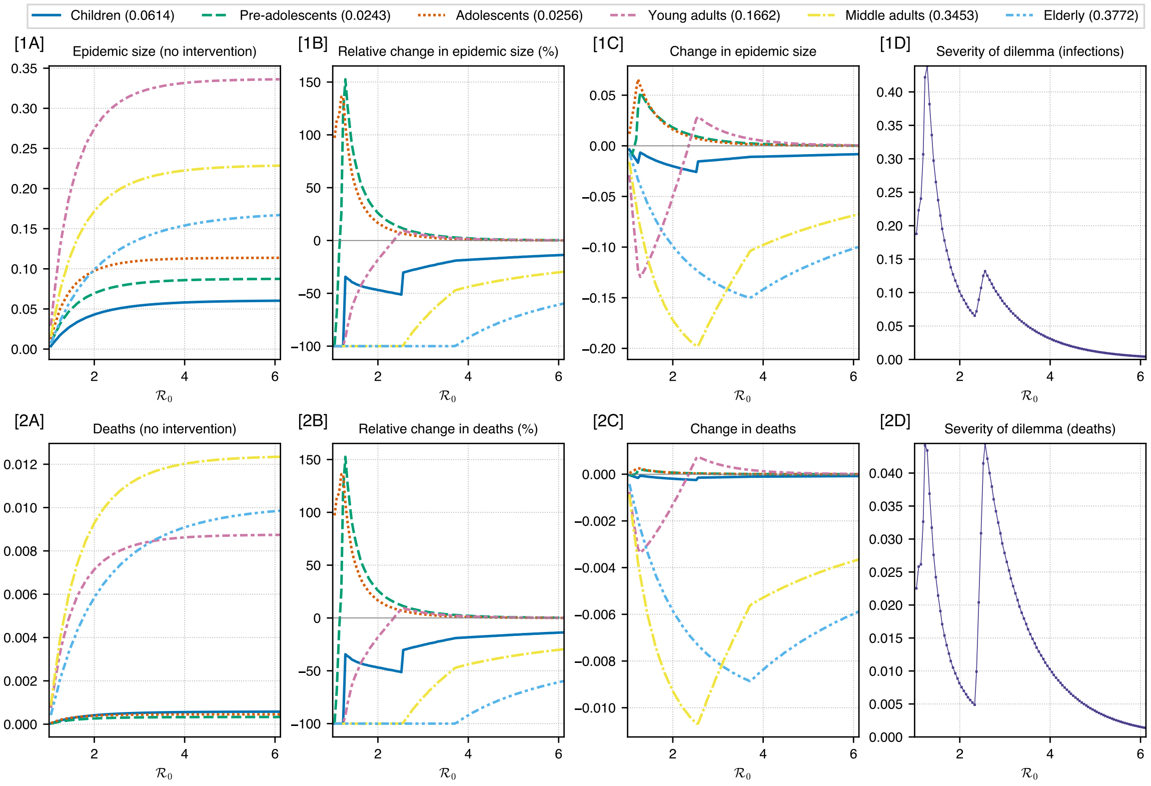


FIG. 4. 2009 flu pandemic: A real-world contact matrix from a sample of the Dutch population is used to determine the effect of optimal intervention on different age groups for a range of $\mathcal{R}_0$ values. Estimates of case fatality rates for 2009 influenza pandemic in Mexico have been used to weight the cost function (for minimising total deaths in the population). Rows (1) and (2) show the plots for infections and deaths respectively. Column (A) shows the epidemic size and deaths if no intervention was performed. Column (B) shows the relative change in epidemic size and deaths under optimal intervention. Column (C) shows the magnitude of change in epidemic size and deaths under optimal intervention. Column (D) shows the severity of the ethical dilemma (see main text for definition) with $\mathcal{R}_0$. The legend for columns (A, B, C) shows age groups and the number in bracket shows the weight assigned to it in the cost function. These weights are proportional to the case fatality rates.

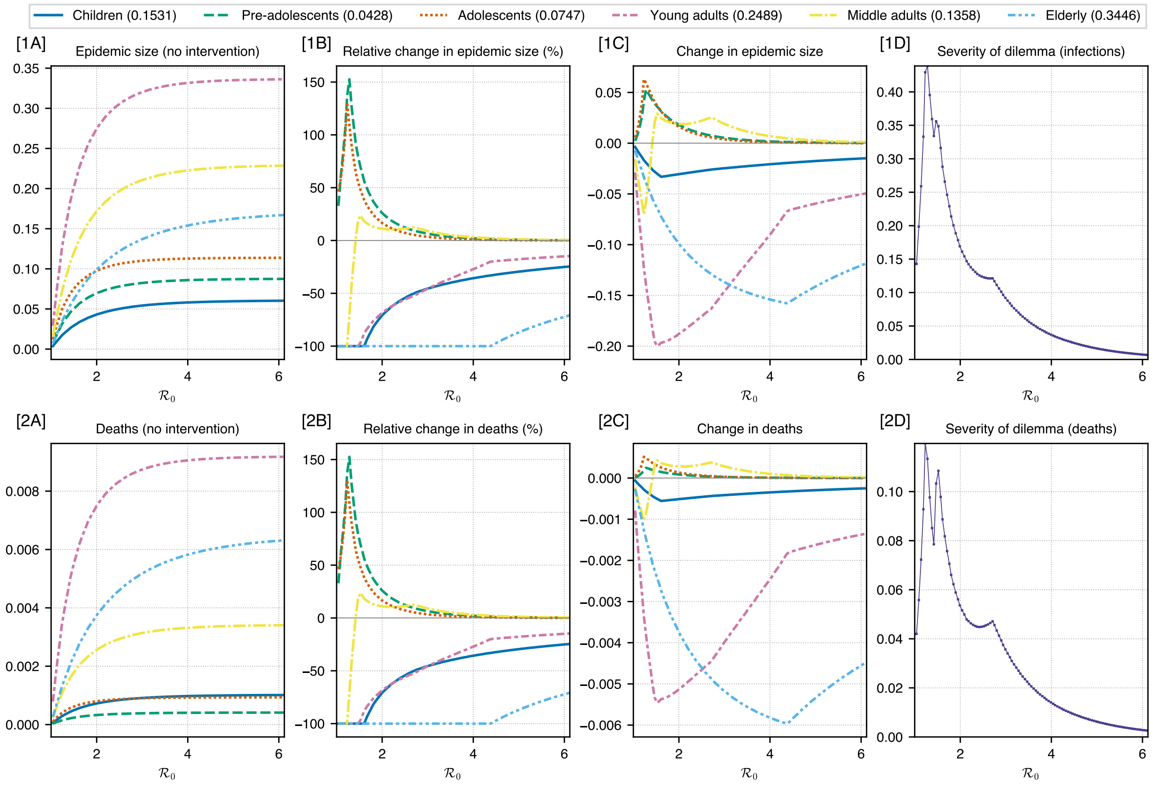


FIG. 5. 1918 flu pandemic: A real-world contact matrix from a sample of the Dutch population is used to determine the effect of optimal intervention on different age groups for a range of $\mathcal{R}_0$ values. Estimates of case fatality rates for the 1918 influenza pandemic have been used to weight the cost function (for minimising total deaths in the population). Rows (1) and (2) show the plots for infections and deaths respectively. Column (A) shows the epidemic size and deaths if no intervention was performed. Column (B) shows the relative change in epidemic size and deaths under optimal intervention. Column (C) shows the magnitude of change in epidemic size and deaths under optimal intervention. Column (D) shows the severity of the ethical dilemma (see main text for definition) with $\mathcal{R}_0$. The legend for columns (A, B, C) shows age groups and the number in bracket shows the weight assigned to it in the cost function. These weights are proportional to the case fatality rates.

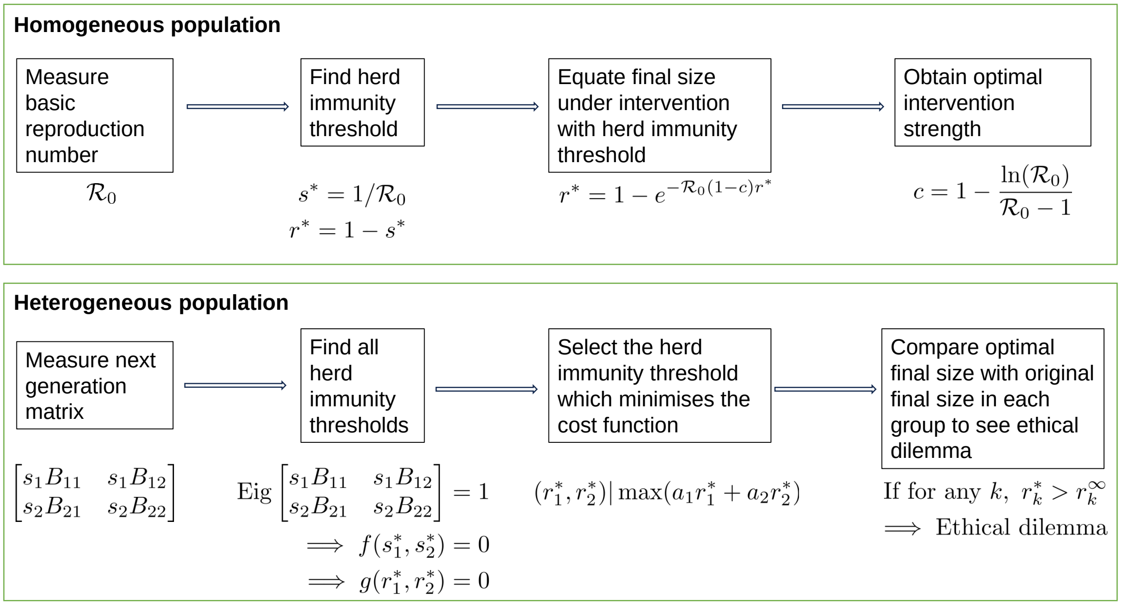


FIG. 6. A schematic description of the optimisation: $s$ and $r$ are the proportion of susceptibles and recovered respectively and with subscripts, they indicate the proportion in a sub-group. 'Eig' refers to the highest eigenvalue. In the heterogeneous case, we only show 2 groups here but this can be generalised to any number of groups.
